# Mediation of calcium-to-phosphorus ratio in the association between kidney stones and bone mineral density in the femoral neck: a cross-sectional study based on the National Health and Nutrition Examination Survey (NHANES)

**DOI:** 10.64898/2026.03.12.26348264

**Authors:** Guihu Liu, Xieyu Wang, Xiaolong Wang, Haibin Zhou, Guangsi Shen

## Abstract

**Background:** Kidney stones, a prevalent urological disorder, are increasingly associated with potential skeletal health issues, including reduced bone mineral density (BMD) and an elevated risk of osteoporosis. However, the underlying mechanisms and subgroup-specific associations have not yet been adequately explored.

**Methods:** This study used data from a nationally representative survey with a weighted complex sampling design. A total of 6,464 participants were enrolled in the study. We performed weighted and unweighted comparative analyses, multivariate linear regression, mediation analysis, and subgroup evaluations to examine the association between kidney stones and BMD of the femoral neck and lumbar spine. Potential mediators, including the systemic immune-inflammation index (SII), estimated glomerular filtration rate (eGFR), and calcium-to-phosphorus (CaP) ratio, were investigated.

**Results:** The presence of kidney stones was significantly associated with lower femoral neck BMD (β =-0.015, p = 0.046) after adjusting for confounding factors. The CaP ratio was identified as a significant mediator (average causal mediation effect [ACME] = 0.00077, p = 0.028), whereas the SII and eGFR did not show significant mediating effects. Stratified analyses revealed stronger associations in participants aged < 50 years and in those without chronic kidney disease (CKD). No significant interactions according to gender were detected.

**Conclusion:** Kidney stones are independently associated with reduced BMD, which is partially mediated by altered calcium-phosphorus homeostasis. These findings highlight the importance of monitoring bone health in patients with kidney stones, particularly in younger and non-CKD populations, and suggest that dietary mineral balance may play a critical role in bone-stone interaction.

## 1. Introduction

Osteoporosis and kidney stones are significant global public health challenges^1,2^. Osteoporosis is a systemic skeletal disorder characterized by reduced bone mass, compromised bone microarchitecture, increased bone fragility, and high risk of fractures^3^. In contrast, kidney stones, particularly those containing calcium, involve complex processes such as solute supersaturation and crystal aggregation in the urine during their formation^4^. Traditionally, osteoporosis and kidney stones have been associated with the skeletal and urinary systems, respectively; however, their interconnections are increasingly recognized for their interconnections^5,6^. Notably, a retrospective cohort study demonstrated that individuals with kidney stones have a lower bone mineral density in the femoral neck and a higher prevalence of osteoporosis^7^. Further research suggests that bone demineralization may contribute to an increased risk of kidney stone formation^8^. This correlation indicated the presence of shared pathophysiological mechanisms between these diseases.

Calcium and phosphorus metabolism disorders are the fundamental links between the pathogenesis of kidney stones and osteoporosis. Hypercalciuria, a common risk factor in individuals with osteoporosis and calcium-based kidney stones^9,10^, not only promotes the formation and growth of calcium oxalate crystals but also adversely affects bone quality^11^. This process is characterized by an imbalance between bone resorption and formation, which is potentially associated with the activation of RANKL and calcium signaling pathways, as well as the stimulation of osteoclastogenesis by inflammatory mediators such as IL-1^5,12,13^. Furthermore, renal insufficiency is another critical pathway that connects kidney stones with decreased bone mineral density^5^. The kidney is pivotal in the regulation of calcium and phosphorus metabolism and activation of vitamin D. Impaired renal function, often evidenced by a reduced estimated glomerular filtration rate, can detrimentally affect bone health through various mechanisms, including disorders of vitamin D and mineral metabolism, systemic inflammation, and oxidative stress. These systemic factors play a role in the pathogenesis of osteoporosis by facilitating osteoclastogenesis and inhibiting osteoblast activity^14,15^. However, notwithstanding these hypothesized mechanisms, the exact relationship between nephrolithiasis and bone mineral density, especially concerning the influence of renal function and biochemical indicators, such as the calcium-to-phosphorus ratio, remains inadequately elucidated in large-scale population studies.

This study used the National Health and Nutrition Examination Survey (NHANES) database and applied weighted survey analysis to explore the following critical questions: (1) Is there an independent association between kidney stone status and bone mineral density (BMD) in the femoral neck and lumbar spine? (2) To what extent does renal function (estimated glomerular filtration rate [eGFR]) and calcium-phosphorus metabolism (calcium-phosphorus ratio, CaP ratio) mediate the relationship between kidney stones and BMD? Additionally, are there variations in this association across different age and sex subgroups? The findings of this study are expected to provide novel epidemiological insights into the underlying comorbidity mechanisms of kidney stones and osteoporosis, with significant clinical implications for the early identification of high-risk populations, development of intervention strategies, and implementation of precise treatments targeting common pathways.

## 2. Material and Methods

### 2.1. Research Design and Data Sources

This cross-sectional study used data from the National Health and Nutrition Examination Survey (NHANES) database, a comprehensive stratified sampling survey administered by the National Center for Health Statistics (NCHS), a branch of the Centers for Disease Control and Prevention (CDC) in the United States. NHANES is conducted biennially and collects cross-sectional data on the demographic characteristics, socioeconomic status, dietary habits, and medical information of American adults and children through surveys, physical examinations, and laboratory analyses. All NHANES data were publicly accessible at www.cdc.gov/nchs/nhanes/. The study protocol was approved by the NCHS Institutional Review Board, and informed consent was obtained from all the participants. An ethics committee waiver exempted the requirement for individual consent forms due to the use of a publicly available database. The work has been reported in line with the STROCSS criteria^16^.

### 2.2. Research Subjects

The study included all adult participants (age≥ 20 years) from the NHANES database from 2007 to 2020 who had undergone assessments of kidney status and BMD. Participants were excluded if they had missing key variables such as kidney stone status, BMD, and eGFR. The final analysis included 6,464 participants (Figure 1), of which 580 were categorized into the kidney stone group and 5,884 into the non-kidney stone group.

**Figure 1.**
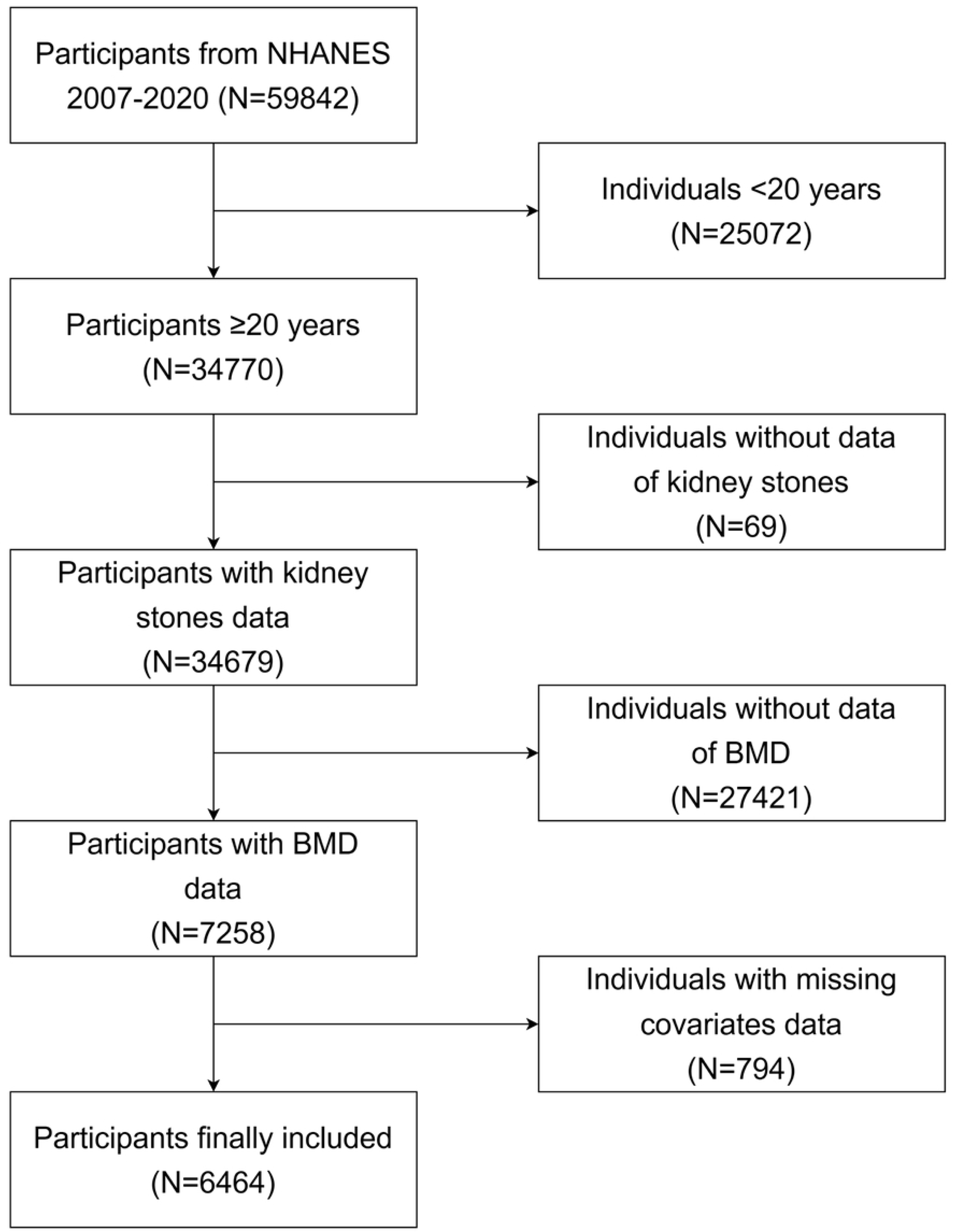
Flow diagram of the study cohort selection.

### 2.3. Variable Definition and Measurement

#### 2.3.1. Main exposure variables

The presence of kidney stones was determined based on self-reported data from the questionnaire, specifically the question, “Have you received a medical diagnosis of kidney stones from a healthcare provider?” Participants who answered affirmatively were classified into the kidney stone group, whereas those who answered negatively were classified into the non-kidney stone group.

#### 2.3.2. Outcome variable

BMD was measured using dual-energy X-ray absorptiometry (DXA), with particular attention to BMD at the femoral neck (FEBMD) and lumbar spine (LBMD) sites. The NHANES database provided BMD data for the first through fourth lumbar vertebrae, and LBMD was calculated as the mean BMD across these four vertebrae. Trained technicians performed DXA measurements according to standardized protocols, ensuring regular calibration of the instruments to maintain data accuracy.

#### 2.3.3. Mediating variable

Renal function was assessed using the eGFR calculated using the Chronic Kidney Disease Epidemiology Collaboration (CKD-EPI) equation, which was specifically adapted for the study’s U.S. population^16^. Calcium and phosphorus metabolism were evaluated using the CaP ratio, which was derived from the serum concentrations of total calcium and phosphorus. Systemic inflammation was quantified using the systemic immune-inflammation index (SII), which was computed as SII = (platelet count × neutrophil count) / lymphocyte count. This index is recognized for its capacity to delineate various inflammatory and immune pathways within the body, thereby providing an accurate reflection of the level of systemic inflammation^17–19^.

#### 2.3.4. Covariate

In accordance with existing literature and theoretical frameworks, we collected and controlled for the following potential confounding variables: demographic factors (including age, sex, race, education level, and poverty-income ratio), anthropometric measures (such as BMI and waist circumference), lifestyle variables (including smoking status and physical activity), laboratory parameters (such as serum total calcium, blood glucose, blood phosphorus, white blood cell count, vitamin D levels, and calcium intake), and comorbid conditions (specifically diabetes and hypertension).

Diabetes was characterized by a medical professional’s diagnosis, glycated hemoglobin levels ≥ 6.5%, fasting blood glucose levels ≥ 7.0 mmol/L, random blood glucose levels ≥ 11.1 mmol/L, and the use of insulin. Hypertension was defined as a medical professional’s diagnosis and a systolic blood pressure of 140 mmHg or higher or diastolic blood pressure of 90 mmHg or higher.

### 2.4. Statistical analysis

The analysis was performed using the R software (version 4.2.2). Initially, weighted demographic characteristics were calculated and baseline features were compared between individuals with and without kidney stones. Categorical variables were analyzed using weighted chi-square tests and continuous variables were evaluated using weighted t-tests or Wilcoxon rank-sum tests. A weighted multiple linear regression model was employed to examine the association between kidney stone status and bone mineral density (specifically, FEBMD and LBMD). Three models were developed: Model 1 (unadjusted), Model 2 (adjusted for age and gender), and Model 3 (adjusted for all covariates). The results are presented as β-estimates with corresponding 95% confidence intervals (CIs). Mediation analysis was conducted to investigate the potential mediating effects of renal function and calcium-phosphorus metabolism on the relationship between kidney stones and BMD.

In this study, we employed the bootstrap method with 500 iterations to evaluate the mediating effects of eGFR, CaP ratio, and SII. This approach facilitated the calculation of the average causal mediation effect (ACME), average direct effect (ADE), total effect, and proportion of mediation effect. We used a restricted cubic spline (RCS) model for visualization to explore the relationship between CaP ratio and bone density. To investigate the potential non-linear relationships between eGFR and BMD, we utilized RCS and B-splines for model fitting and visualization of the predicted outcomes. Subgroup analyses were conducted to examine potential variations in associations across diverse populations. The stratification criteria included age (<50 vs. ≥50 years), sex (male vs. female), diabetes status (presence vs. absence), chronic kidney disease (chronic kidney disease [CKD], defined as eGFR < 60 ml/min/1.73 m² vs. eGFR ≥ 60 ml/min/1.73 m²), and obesity (body mass index [BMI] ≥ 30 kg/m ² vs. BMI < 30 kg/m ²). Interaction terms, such as kidney stones × sex, were incorporated into the model to assess the significance of the interactions. A sensitivity analysis was performed to address the potential impact of severe renal insufficiency by limiting the analysis to patients with eGFR ≥ 30 mL/min/1.73 m² and refitting the primary model. Multicollinearity diagnostics were performed for all the regression models. All statistical tests were two-sided, and statistical significance was set at P < 0.05.

## 3. Results

### 3.1. Characteristics of the Study Population

The study included 6,464 participants, with 580 individuals (9%) in the kidney stone group and 5,884 individuals (91%) in the non-kidney stone group. Table 1 presents the baseline characteristics of the participants before and after weighing. Comparative analyses indicated that participants with kidney stones were older (mean age 52.3 years versus 47 years, p < 0.001) and had a higher proportion of males (61.2% vs. 47.7%, p < 0.001). The FEBMD was significantly lower in the kidney stone group than in the non-stone group (0.82 versus 0.84 g/cm², p = 0.035), whereas no significant difference was observed in the lumbar spine bone mineral density. Renal function, as assessed by eGFR, was significantly reduced in the kidney stone group (94.96 versus 99.72 ml/min/1.73m², p < 0.001). Additionally, the CaP ratio (2.03 vs. 1.98, p = 0.001) and SII (572.63 vs. 533.41, p = 0.039) were significantly higher in the kidney stone group. Furthermore, the prevalence of diabetes (18.5% vs. 8%, p < 0.001) and hypertension (49.4% vs. 30.7%, p < 0.001) were higher in the kidney stone group.

**Table 1.** Weighted characteristics of the study population grouped by kidney stone status.

|  | Kidney stone<br>(N=580) | No kidney stone<br>(N=5884) | p-value |
| --- | --- | --- | --- |
| Age, n (%) |  |  | <0.001 |
| 20-30 | 39 (8.9) | 1084 (20.3) |  |
| 31-40 | 83 (13.7) | 1036 (18.3) |  |
| 41-50 | 130 (27.6) | 1383 (26.6) |  |
| 51-60 | 118 (24.7) | 1042 (18.9) |  |
| 61-70 | 120 (14.9) | 811 (10.0) |  |
| 71-80 | 59 (6.7) | 363 (4.0) |  |
| ≥80 | 31 (5.3) | 165 (1.9) |  |
| Gender, n (%) |  |  | <0.001 |
| Male | 347 (61.2) | 2831 (47.7) |  |
| Female | 233 (38.8) | 3053 (52.3) |  |
| Race, n (%) |  |  | <0.001 |
| Non-Hispanic White | 331 (78.7) | 2619 (67.7) |  |
| Non-Hispanic black | 68 (5.5) | 1055 (10.6) |  |
| Mexican American | 87 (5.9) | 1148 (9.4) |  |
| Other | 94 (9.8) | 1062 (12.3) |  |
| Education, n (%) |  |  | 0.957 |
| Below high school | 160 (17.7) | 1584 (17.8) |  |
| High school or above | 420 (72.4) | 4300 (82.2) |  |
| PIR, n (%) |  |  | 0.601 |
| Poor | 119 (12.7) | 1156 (13.7) |  |
| Not poor | 461 (87.3) | 4728 (86.3) |  |
| Physical activities, n (%) |  |  | 0.015 |
| No | 486 (79.6) | 4521 (72.5) |  |
| Yes | 94 (20.4) | 1363 (27.5) |  |
| Alcohol use, n (%) |  |  | <0.001 |
| No | 536 (92.3) | 5237 (86.4) |  |
| Yes | 44 (7.6) | 647 (13.6) |  |
| Smoking status, n (%) |  |  | 0.001 |
| Never | 285 (49.9) | 3291 (56.4) |  |
| Former | 178 (29.5) | 1258 (21.7) |  |
| Current | 117 (20.6) | 1335 (22.0) |  |
| Diabetes, n (%) |  |  | <0.001 |
| No | 450 (81.5) | 5191 (92.0) |  |
| Yes | 130 (18.5) | 693 (8.0) |  |
| Hypertension, n (%) |  |  | <0.001 |
| No | 264 (50.6) | 3819 (69.3) |  |
| Yes | 316 (49.4) | 2065 (30.7) |  |
| FEBMD (g/cm<sup>2</sup>) | 0.82 (0.14) | 0.84 (0.15) | 0.035 |
| LBMD (g/cm<sup>2</sup>) | 1.03 (0.15) | 1.03 (0.14) | 0.987 |
| CaP ratio | 2.03 (0.31) | 1.98 (0.32) | 0.001 |
| eGFR (ml/min/1.73m<sup>2</sup>) | 94.96 (20.78) | 99.72 (20.96) | <0.001 |
| SII | 572.63 (646.8) | 533.41 (309.97) | 0.039 |
| Vitamin D (nmol/L) | 63.25 (24.95) | 65.06 (26.11) | 0.243 |
| Calcium intake (mg) | 888.21 (496.45) | 901.83 (528.46) | 0.653 |
| Total Calcium (mmol/L) | 2.35 (0.09) | 2.36 (0.09) | 0.19 |
| Phosphorus (mmol/L) | 1.18 (0.18) | 1.22 (0.18) | <0.001 |
| WBC (mmol/L) | 7.40 (2.80) | 7.11 (2.18) | 0.028 |
| Waist circumference (cm) | 101.44 (15.04) | 95.62 (14.21) | <0.001 |
| BMI (kg/m<sup>2</sup>) | 29.25 (5.82) | 27.7 (5.54) | <0.001 |

Mean (SE) for continuous variables: the p-value was calculated by the weighted one-way analysis of variance. n (%) for categorical variables: the p-value was calculated by the weighted chi-square test. Abbreviation: PIR, ratio of family income to poverty; FEBMD, femoral neck bone mineral density; LBMD, lumbar bone mineral density; CaP ratio, calcium-phosphorus ratio; eGFR, estimated glomerular filtration rate; SII, systemic immune-inflammation index; WBC, white blood cell; BMI, body mass index. p<0.05 is statistically significant.

### 3.2. Associative Analysis of Kidney Stones and BMD

After adjusting for various confounding variables, including age, sex, waist circumference, eGFR, vitamin D levels, diabetes, and hypertension, the weighted multiple linear regression analysis identified a significant inverse association between the presence of kidney stones and reduced FEBMD (β =-0.015, 95% CI:-0.030 to - 0.001, p = 0.046) (Table 2). In contrast, the relationship between kidney stones and LBMD was not statistically significant (β =-0.007, p = 0.289). Additionally, the above conclusion holds and is more significant after adding the interaction terms.

**Table 2.**
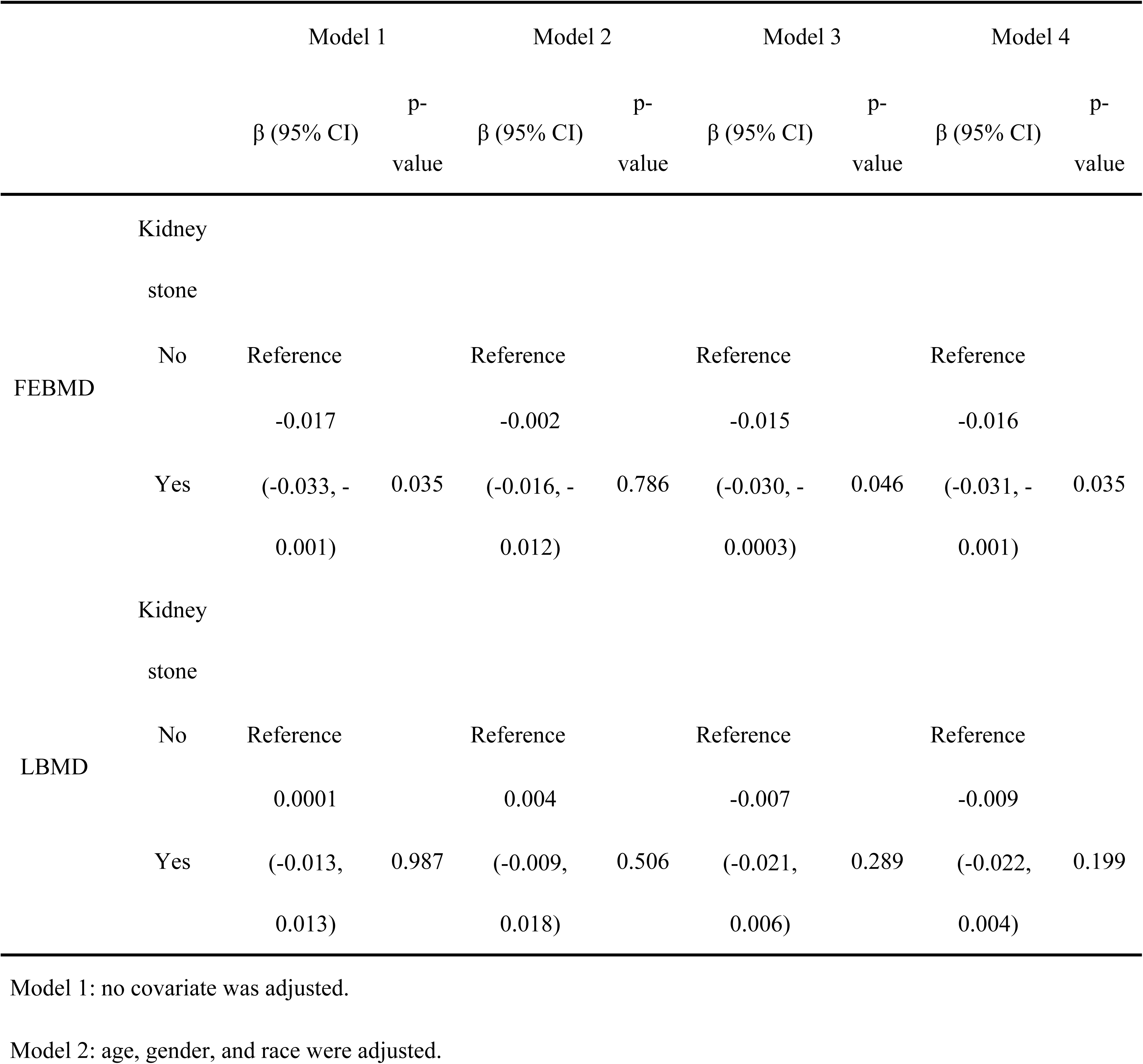
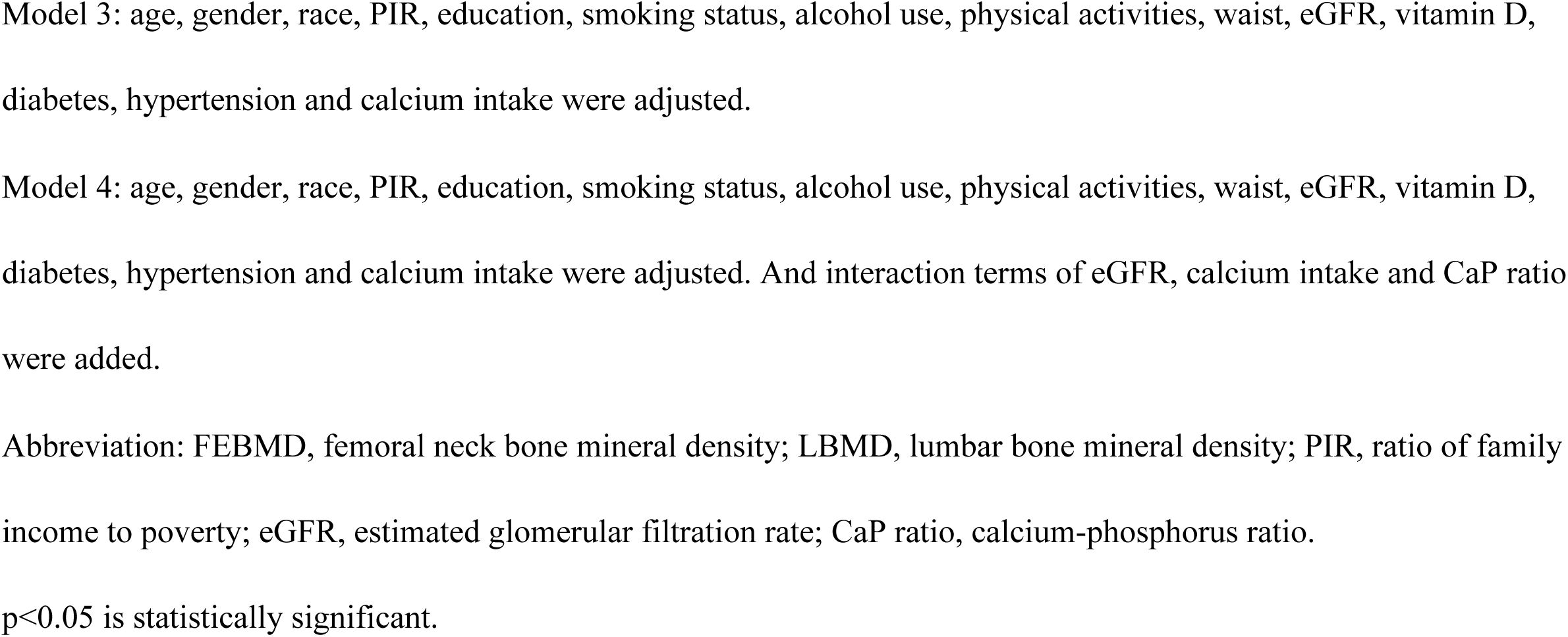
Relationship between bone mineral density and kidney stone status in participants aged ≥ 20 years.

### 3.3. Subgroup Analysis and Interaction

Analysis stratified by age indicated a more pronounced negative correlation between kidney stones and FEBMD in individuals younger than 50 years (β =-0.025, 95% CI:-0.048 to-0.002, p = 0.039) (Figure 2). Conversely, in individuals aged ≥ 50 years, this correlation was attenuated and did not reach statistical significance (β =-0.007, p = 0.393). An interaction test suggested that age might modulate the association between kidney stones and BMD, with a borderline significant interaction p-value of 0.098.

**Figure 2.**
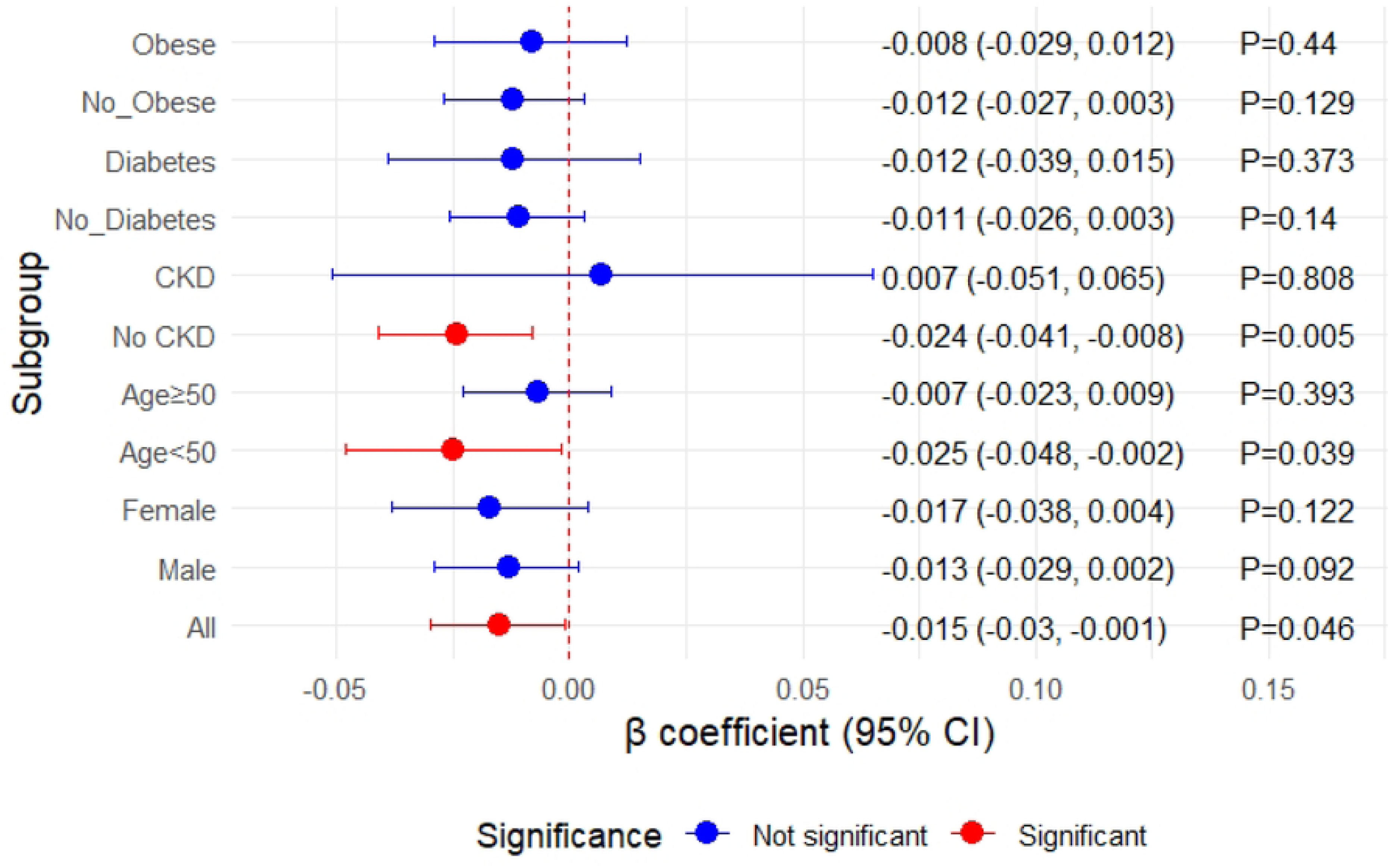
Subgroup analysis of the effect of kidney stones on BMD of the femoral neck. CKD, chronic kidney disease; CI, confidence interval; BMD, bone mineral density.

A gender-stratified analysis indicated a non-significant inverse relationship between kidney stones and FEBMD in both males (β =-0.013, p = 0.092) and females (β =-0.017, p = 0.122), with no significant interaction by gender (P = 0.798). Further stratification based on renal function revealed a significant association between kidney stones and FEBMD among participants without CKD (β =-0.024, P = 0.005), whereas no significant association was observed in those with CKD (β = 0.007, P = 0.808), and the interaction was not significant (P = 0.318). When stratified by diabetes status, the association between kidney stones and FEBMD was more evident in non-diabetic individuals (β =-0.011, P = 0.140) than in diabetic patients (β =-0.012, P = 0.373), although the association was not statistically significant and the interaction was non-significant (P = 0.918). Stratification by BMI demonstrated a non-significant association between kidney stones and FEBMD in both the non-obese (β =-0.012, P = 0.129) and obese (β =-0.008, P = 0.440) groups, with a non-significant interaction (P = 0.849).

### 3.4. Mediating Effect Analysis

Mediation analysis identified a significant mediating effect of CaP ratio on the relationship between kidney stones and FEBMD (Table 3). The ACME was 0.00078 (95% CI: 0.000009–0.001603, p = 0.048), accounting for 14.7% of the total effect. These results indicate that the adverse effect of kidney stones on bone mineral density is partially mediated by changes in calcium and phosphorus metabolism.

**Table 3.** Results of mediation effect analysis.

| Mediating<br>variable | ACME | 95% CI | p-value | ADE | Total effect | Intermediary<br>Ratio |
| --- | --- | --- | --- | --- | --- | --- |
| SII | -0.00035 | (-0.00076, 0.00011) | 0.160 | -0.00355 | -0.00390 | 9.0 |
| eGFR | -0.00066 | (-0.00279, 0.00135) | 0.472 | -0.00549 | -0.00615 | 10.8 |
| CaP ratio | 0.00077 | (0.00008, 0.00170) | 0.028 | -0.00588 | -0.00510 | -15.2 |

p<0.05 is statistically significant.
Abbreviation: ACME, average causal mediation effect; CI, confidence interval; ADE, average direct effect; eGFR, estimated glomerular filtration rate; SII, systemic immune-inflammation index; CaP ratio, calcium-phosphorus ratio.

### 3.5. Visualization of the Dose Relationship between CaP Ratio and BMD

The dose-response relationship curve plotted using a restricted cubic spline model revealed a significant non-linear association between the CaP ratio and femoral neck bone density, and this association pattern varied by sex (Figure 3). In male participants, the relationship between the CaP ratio and BMD exhibited a slight U-shaped curve trend, whereas in female participants, we observed a more complex non-linear relationship resembling a non-monotonic curve that first decreased and then increased.

**Figure 3.**
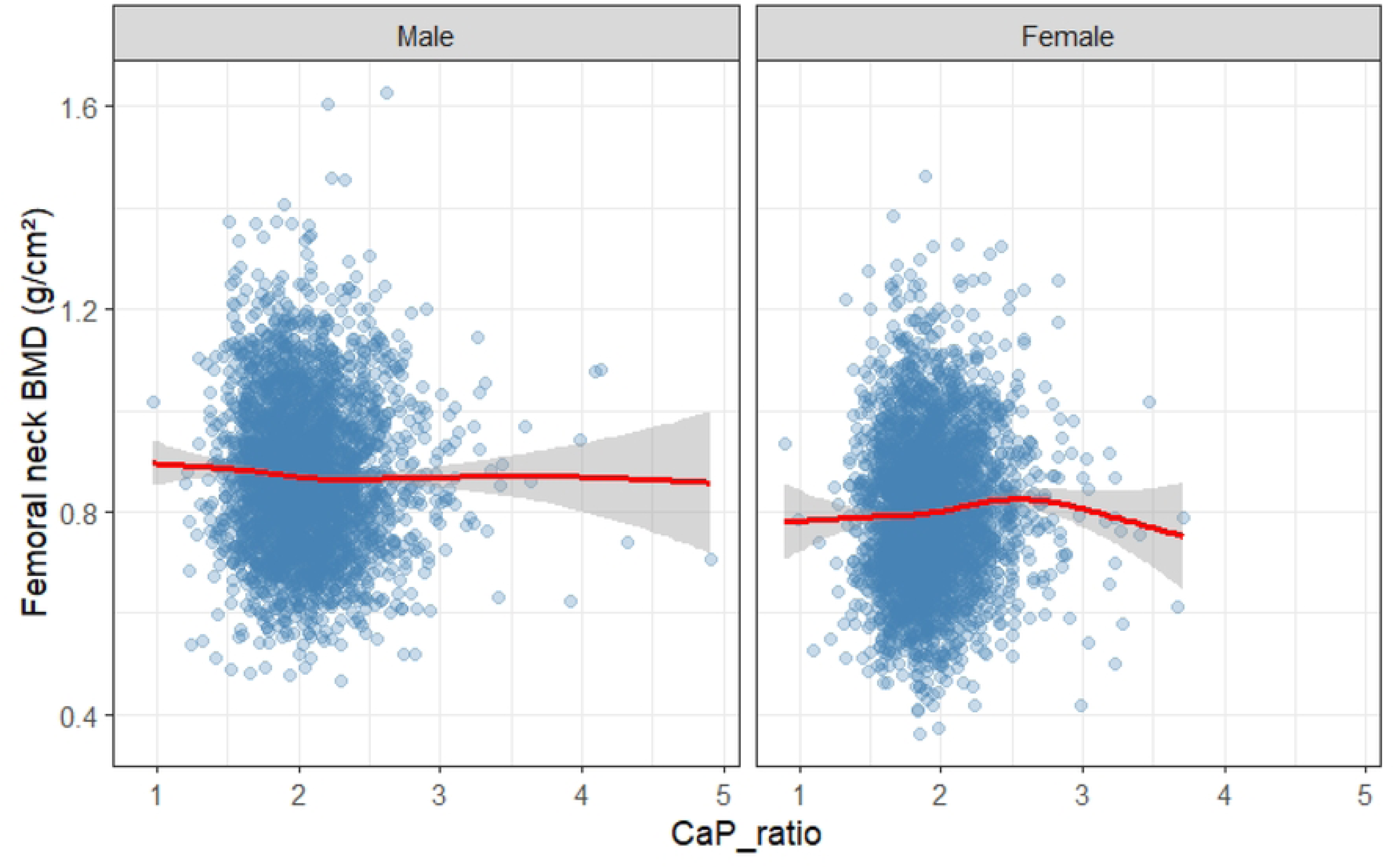
The non-linear relationship between CaP ratio and femoral neck bone density. The red line represents the trend of dose variation, while the gray area represents the 95% confidence interval. BMD, bone mineral density.

### 3.6. Exploration of Non-linear Relationships

RCS and B-splines were used to assess the nonlinear relationship between eGFR and FEBMD (Figure 4). Both methods demonstrated a non-linear association between the eGFR and BMD. Specifically, at lower eGFR levels, particularly below 60 ml/min/1.73m², a marked decline in BMD was observed with decreasing eGFR. In contrast, when eGFR exceeded 90 ml/min/1.73m², the relationship between BMD and eGFR appeared to reach a plateau.

**Figure 4.**
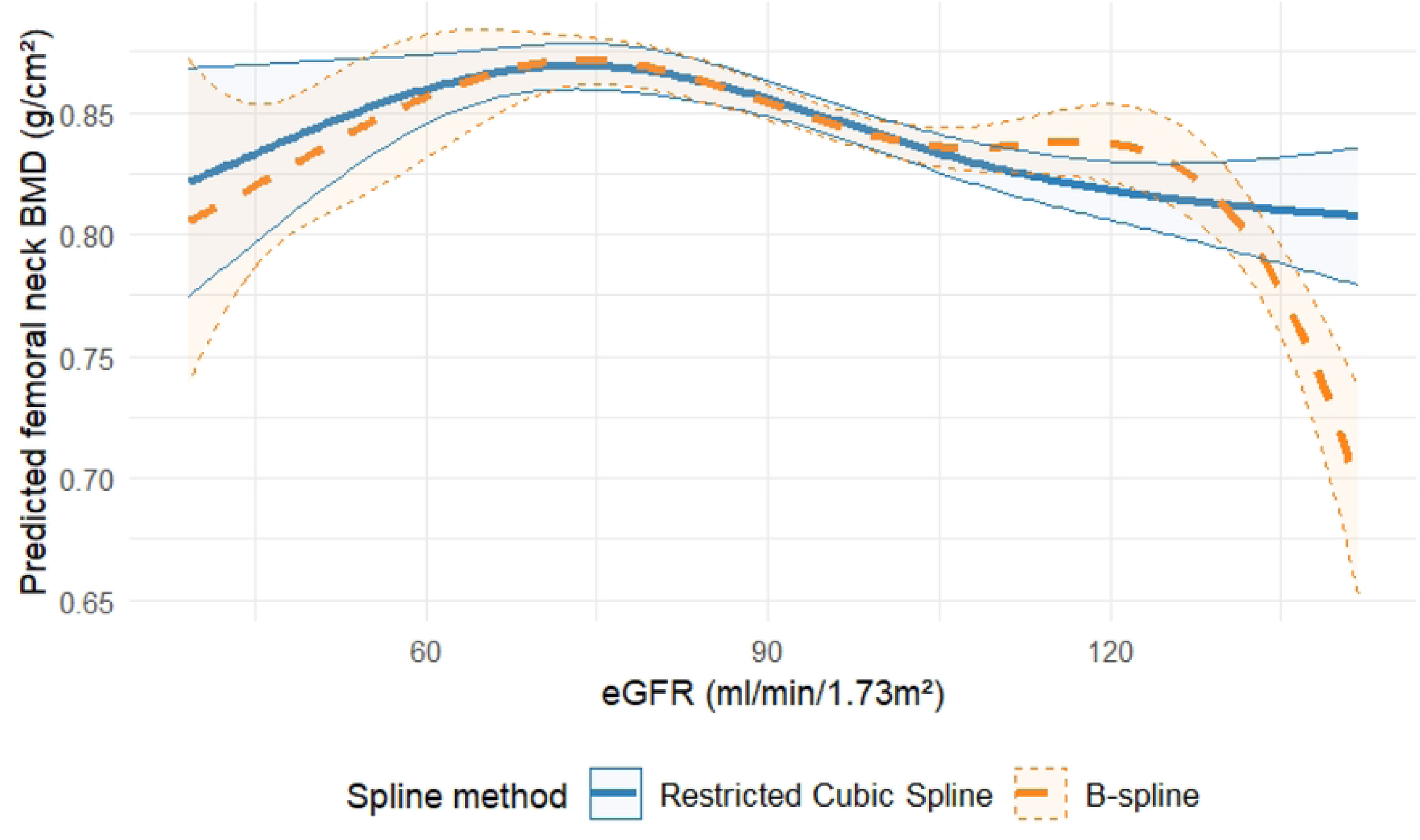
Nonlinear relationship between eGFR and BMD of the femoral neck: restricted cubic spline vs B-spline. BMD, bone mineral density; eGFR, estimated glomerular filtration rate.

### 3.7. Sensitivity Analysis

To address the potential impact of severe renal insufficiency, a sensitivity analysis was conducted by restricting the study population to patients with an eGFR of ≥ 30 ml/min/1.73m² (n = 6,352). Following the recalibration of the model, the inverse association between kidney stones and FEBMD remained statistically significant (β =-0.015, 95% CI:-0.030 to-0.001, p = 0.045), consistent with the findings of the primary analysis, thereby reinforcing the robustness of the study results.

### 3.8. Multicollinearity Test

A multicollinearity assessment of all regression models indicated variance inflation factors (VIFs) below five, with a maximum VIF of 3.09, suggesting that significant multicollinearity issues were not present.

## 4. Discussion

By leveraging data from the NHANES, a comprehensive and nationally representative database, this study conducted an in-depth analysis of the relationship between kidney stones and BMD as well as the underlying mechanisms involved. The principal findings of this study were as follows: (1) individuals with kidney stones demonstrated significantly lower BMD at the femoral neck, independent of various traditional risk factors. (2) Alterations in calcium-phosphorus metabolism, as indicated by the CaP ratio, significantly mediated the relationship between kidney stones and BMD, accounting for approximately 15% of the total effect. (3) The observed association was more pronounced among individuals aged < 50 years and those without CKD. (4) A complex nonlinear relationship between renal function, as measured by eGFR and BMD, was identified. These findings provide novel epidemiological insights and mechanistic perspectives to understand the intrinsic link between kidney stones and osteoporosis, both of which are prevalent.

### 4.1. Mechanistic Explanation of the Association between Kidney Stones and BMD

This study demonstrated a significant reduction in FEBMD among patients with kidney stones (β =-0.015, p = 0.046), corroborating the findings of previous research^20,21^. The primary mechanism underlying this reduction appears to involve the disruption of calcium and phosphorus metabolism. Our mediation analysis identified the CaP ratio as a critical mediator in the relationship between kidney stones and BMD (ACME = 0.00078, p = 0.048). Individuals with kidney stones, particularly those with calcium-based stones, frequently exhibit hypercalciuria, which is a significant risk factor for stone formation and may contribute to calcium depletion from bones^4^. The etiology of hypercalciuria may originate from increased intestinal calcium absorption or decreased renal calcium reabsorption. However, emerging evidence indicates that enhanced bone resorption significantly contributes to this condition^22^. A study conducted in male patients with hypercalciuric kidney stones in France revealed that fasting hypercalciuria was the only biological factor associated with reduced BMD. Notably, when the fasting calcium-to-creatinine ratio exceeded 0.25 mmol/mmol, the likelihood of low BMD increased by 3.8 times^23^. This finding resonates with our observation that the CaP ratio was elevated in the kidney stone cohort (2.03 vs. 1.98, p = 0.001).

Decline in renal function is recognized as a critical area for investigation^24^. This study identified a significant difference in eGFR between individuals with kidney stones and those without (94.96 vs. 99.72 ml/min/1.73m², p < 0.001), and a non-linear relationship was observed between eGFR and BMD. Additionally, stratified analysis demonstrated a stronger association between kidney stones and BMD in patients without CKD (P _noCKD_ = 0.005 vs. P_CKD_ = 0.808). Patients with CKD frequently experience mineral and bone disorders characterized by disturbances in calcium and phosphorus metabolism, elevated parathyroid hormone (PTH) levels, and abnormalities in vitamin D metabolism^14^. CKD progression results in decreased renal 1α-hydroxylase activity, leading to reduced synthesis of 1, 25-dihydroxyvitamin D3 (calcitriol) synthesis, diminished intestinal calcium absorption, secondary hyperparathyroidism, increased bone resorption, and subsequent bone loss^14,25,26^. Systemic inflammation may be involved in this process. The study revealed a significantly elevated SII in the kidney stone cohort compared with that in the non-stone cohort (572.63 vs. 533.41, p = 0.039). Although the mediating effect was not statistically significant (p = 0.116), it suggests the potential involvement of inflammatory processes. Previous studies have identified increased levels of inflammatory cytokines such as IL-1 and TNF-α in individuals with kidney stones. These cytokines not only contribute to stone formation^27^ but may also promote osteoclastogenesis and enhance bone resorption by activating the RANKL/RANK/OPG pathway^28^. Moreover, osteopontin (OPN), a critical bone matrix protein found in both bone tissue and kidney stones, may influence the regulation of bone metabolism and formation of kidney stones^29^.

### 4.2. Possible Explanations for Age Heterogeneity

A significant finding of this study was the age-related modification in the correlation between kidney stones and BMD. In individuals aged < 50 years, a stronger association was identified between kidney stones and FEBMD (β =-0.025, p = 0.039). In contrast, this association diminished and lost statistical significance in individuals aged ≥ 50 years (p = 0.098). This phenomenon may be explained by the fact that in younger and middle-aged individuals, kidney stone formation is more likely to be linked to metabolic factors such as idiopathic hypercalciuria, which also affects bone health. In contrast, osteoporosis in the elderly is primarily driven by age-related factors, including declining sex hormone levels and age-related bone loss, which may obscure the independent effect of kidney stones on bone health.

Additionally, prevalent comorbidities, such as arthritis and degenerative diseases, along with medication use, including diuretics and glucocorticoids, among older adults, may introduce confounding variables into this relationship. These risk factors are associated with kidney stone formation and bone density^30,31^.

Our research challenges this common belief by demonstrating a stronger link between kidney stones and bone loss in premenopausal women and middle-aged men. This finding supports a cohort study that found the highest fracture risk from urolithiasis in women aged 30-39 and men aged 10-19^32^. Conversely, another study found no significant association between changes in bone mineral density and urinary stones in postmenopausal women^33^, contradicting the idea that postmenopausal women are more prone to bone loss due to estrogen deficiency^34^. These results suggest that, in younger populations, kidney stones may be an important indicator of systemic mineral metabolism issues, with negative effects on bone health appearing early.

### 4.3. Thoughts on Site-specific Differences

This study identified a significant correlation between kidney stones and FEBMD; however, no such association was found with LBMD. This site-specific discrepancy may be attributed to the differences in bone turnover rates and tissue structures across various skeletal regions. The femoral neck is primarily composed of cortical bone, whereas the lumbar spine mainly consists of cancellous bone. Cortical and cancellous bones exhibit differences in metabolic activity, responses to mechanical stimuli, and hormonal regulation^35^. Previous studies have demonstrated that individuals with kidney stones experience more pronounced bone loss in the femoral region than in the lumbar spine^36^, which is consistent with our findings. These observations suggest that kidney stone-related bone loss follows a distinct pattern rather than exerting a uniform effect on the skeletal system.

### 4.4. Clinical Significance and Translational Value

The results of the present study have several clinical implications. Individuals with kidney stones, particularly middle-aged or younger individuals, should be regarded as a high-risk group for developing osteoporosis. Clinicians should assess bone mineral density and fracture risk, particularly in patients with recurrent kidney stones.

Addressing disorders of calcium and phosphorus metabolism is essential for preventing stone recurrence and bone loss. In patients with hypercalciuria, thiazide diuretics may be used to decrease urinary calcium excretion, whereas citrate preparations may be beneficial in correcting hypocitraturia, thereby enhancing bone mineral density^37,38^. Moreover, a more thorough evaluation of the relationship between renal function and bone health is warranted. Even among patients with kidney stones with ostensibly normal estimated glomerular filtration rates (eGFR >90 ml/min/1.73 m² as observed in this study), early renal function impairment or tubular dysfunction may already be present, potentially affecting mineral metabolism and indirectly compromising bone health.

### 4.5. Research Advantages and Limitations

This study has several notable strengths, including the use of an extensive NHANES dataset, which is characterized by a large and nationally representative sample size. This study employed a weighted analysis approach to address the complex sampling design and was thoroughly adjusted for multiple potential confounders. Furthermore, this study incorporated mediation and stratified analyses to explore underlying mechanisms and potential effect modifications. However, this study had several limitations. First, the cross-sectional design precludes the establishment of causal relationships and does not fully exclude the possibility of reverse causality such as the influence of abnormal bone metabolism on stone formation. Second, the reliance on self-reported diagnoses of kidney stones introduces the potential for misclassification. Third, despite extensive adjustment for confounders, the possibility of residual confounding remains, particularly due to the omission of crucial bone metabolism indicators such as PTH and bone turnover markers. Finally, while mediation analysis relies on conventional regression techniques with bootstrap estimation of standard errors, caution is advised when interpreting the causal mediation effects.

## 5. Conclusions

This study confirmed an independent association between nephrolithiasis and reduced bone mineral density of the femoral neck, particularly in younger populations.

Alterations in calcium and phosphorus metabolism were identified as the key mediating factors in this relationship. These findings highlight the importance of considering kidney stones as relevant markers of bone health. Therefore, it is recommended to conduct bone mineral density screening and implement personalized interventions.

## Declarations

## Data Availability

Data are open access and can be downloaded at: https://wwwn.cdc.gov/Nchs/Nhanes/.

## Acknowledgements

The authors would like to express sincere appreciation to all the participants in the National Health and Nutrition Examination Survey (NHANES) and the researchers at the National Center for Health Statistics (NCHS), who collected and managed the NHANES data and also provided the data for public use. Additionally, authors would like to thank *Home for Researchers* team (https://www.home-for-researchers.com).

## Authors’ contributions

Guihu Liu: Data curation, Formal analysis, and Writing original draft; Xieyu Wang: Writing - Reviewing and Editing; Xiaolong Wang: Writing - Reviewing and Editing; Haibin Zhou: Writing - Reviewing and Editing, Funding; Guangsi Shen: Conceptualization, Writing original draft, Writing - review & editing, and Validation.

## Data availability

No datasets were generated or analysed during the current study.

## Ethics approval and consent to participate

The study protocol was approved by the NHANES Institutional Review Board, and was performed in accordance with the Declaration of Helsinki, with all NHANES participants providing signed informed consent.

## Consent for publication

Not applicable.

